# Lifestyle activities in mid-life contribute to cognitive reserve in middle-aged individuals at risk for late-life Alzheimer’s disease, independent of education and occupation

**DOI:** 10.1101/2023.07.04.23292189

**Authors:** Feng Deng, Sandra El-Sherbiny, Maria-Eleni Dounavi, Karen Ritchie, Graciela Muniz-Terrera, Paresh Malhotra, Craig W Ritchie, Brian Lawlor, Lorina Naci

**Author notes:** **Corresponding author:** Lorina Naci, School of Psychology, Trinity College Institute of Neuroscience Global Brain Health Institute, Trinity College Dublin Dublin, Ireland.

## Abstract

It is now acknowledged that Alzheimer’s disease (AD) neuropathology starts decades before the onset of clinical symptoms, but it remains unknown whether modifiable lifestyle factors can protect against these incipient AD processes, early, in mid-life. Cognitive reserve can explain cognitive preservation in some older adults despite ageing or dementia symptoms, but it is not known whether it can protect against neurodegeneration in mid-life. We asked whether modifiable lifestyle activities contribute to cognitive reserve in mid-life, and whether it can offset the risk of AD in individuals who are cognitively healthy. Cognition, structural, and functional brain health measures were assessed at baseline and two years follow-up, in a cohort of middle-aged participants (N = 210; 40–59 years). Mid-life activities were measured using the Lifetime of Experiences Questionnaire. We assessed the impact of lifestyle activities and known risk factors for sporadic late-onset AD (i.e., the Cardiovascular Risk Factors Aging and Dementia [CAIDE] score) on measures of cognition and brain health. Multivariable linear regression found that mid-life activities made a unique contribution to cognition, independent of education and occupation. Crucially, mid-life activities moderated the relationship between cognitive ability (verbal and visuospatial functions, and conjunctive short-term memory binding) and brain health. Cognitive ability of people with higher mid-life activities, particularly those with high dementia risk scores, was less dependent on their brain functional architecture. Impaired visuospatial function is one of the earliest cognitive deficits in AD and has previously been associated with increased AD risk in this cohort. Additionally, conjunctive memory functions have been found impaired in the pre-symptomatic stages of AD. These findings suggest that modifiable activities contribute uniquely to cognitive reserve in midlife, and may offset the risk of AD. The modifiability of these activities supports their targeting by public health initiatives aimed at dementia prevention.

## 1 Introduction

Dementia is a growing epidemic that presents profound challenges to health care systems, families, and societies throughout the world. Alzheimer’s disease (AD), the most common etiology of dementia, is characterized by relentless neurodegeneration and accelerated cognitive decline in the years following presentation of clinical symptoms. It is now accepted that AD neuropathology is present decades before symptoms appear (Jack et al., 2013; Ritchie et al., 2015).

Growing evidence suggests that up to 40% of all dementia cases are associated with known lifestyle-related modifiable risk factors, such as alcohol consumption, obesity and hypertension, among others (Livingston et al., 2020). As exposure to most of these risk factors begins decades before dementia onset, interventions must be implemented in mid-life (Gottesman et al., 2017; Irwin et al., 2018; Lachman et al., 2015). Mid-life, thus, presents a critical and unique window for disease-altering interventions, before the manifestation of substantial brain damage. As a multidimensional construct, lifestyle has a multipronged impact on cognition and the brain. By contrast to aforementioned risk factors, a number of other lifestyle factors such as education, activities that stimulate the brain (e.g., as socialising, reading, learning new skills, regular exercise, etc.), and occupational attainment have been associated with preservation of cognitive function in older adults (Chan et al., 2018) and reduced symptom severity in Alzheimer’s disease (Dekhtyar et al., 2019; Livingston et al., 2020; Wang et al., 2017), a phenomenon known as “cognitive reserve” (Stern et al., 2020). These lifestyle factors are thought to explain why there is considerable variability in the degree of cognitive impairment in late-onset AD (Bennett et al., 2003; Ewers et al., 2013), even when accounting for key pathologies, including beta-amyloid (Aβ) and pathological tau (Franzmeier et al., 2020; Jack & Holtzman, 2013). While epidemiological evidence strongly suggests that education and occupation contribute to cognitive resilience in late life (Deary 2005), there is a renewed interest in the additional contribution of other activities undertaken in mid-life, given their potential modifiability in mid-life. The early accumulation of cognitive reserve in mid-life, decades before dementia onset, may delay, reduce symptom severity, and ultimately prevent late-life AD, but remains poorly understood.

A rigorous definition of cognitive reserve predicts not only that lifestyle factors will be associated with better cognition, but also that they will moderate the relationship between brain health and cognition (Brayne et al., 2010; Song et al., 2022; Stern et al., 2020). In other words, the cognitive abilities of individuals with high reserve should be de-coupled from typical measures of brain health, such as grey matter volume (Nilsson & Lövdén, 2018; Stern, 2017), but rather be realized through compensatory brain mechanisms. In support of this aspect of cognitive reserve, Chan et al. (2018) found that physically, socially, and cognitively stimulating lifestyle activities undertaken in midlife significantly moderated the relationship between total grey matter volume (GMV) and fluid intelligence in healthy older adults (aged 66–88 years). Older people with higher levels of engagement in stimulating activities did not show an association between cognition and GMV. Their cognitive ability was maintained regardless of decrements in brain structure, in stark contrast to those with low engagement levels, who showed a significant association between lower GMV and lower cognition, in keeping with expectations. This and other studies have established that stimulating activities contribute to cognitive reserve in late life, but it remains unknown whether this form of cognitive reserve is present in mid-life and, particularly in individuals at risk for AD. Answering this question is crucial for identifying early interventions for building cognitive reserve, which may delay the onset and reduce symptom severity in late-life Alzheimer’s disease.

We have previously shown that more frequent engagement in physically, socially, and intellectually stimulating activities undertaken in midlife were significantly associated with better cognition in midlife (Heneghan et al., 2023), independently of education and occupational attainment. However, it remains unknown whether stimulating lifestyle activities moderate the relationship between cognition and measures of brain health in midlife, especially in at-risk individuals. To address this gap, we asked (a) whether stimulating lifestyle activities moderate the relationship between cognition and measures of brain health in midlife, and (b) whether the moderation effects are present in individuals at high risk for late-life AD. We used a composite dementia risk score, i.e., the Cardiovascular Risk Factors, Aging, and Incidence of Dementia (CAIDE) (Kivipelto et al., 2006), to define the at-risk individuals, as it has been used to select at-risk individuals in large lifestyle intervention trails in older adults (> 60 years) (Ngandu et al., 2015). By doing so, we could relate our results to previous studies (Kivipelto et al., 2018).

We addressed these questions in a cognitively healthy middle-aged cohort (N = 210; aged 40–59 years) from the Imperial College London site of the PREVENT-Dementia research programme (https://preventdementia.co.uk/), assessed cross-sectionally and at two-years follow-up. Participants underwent extensive cognitive assessment and multimodal Magnetic Resonance Imaging (MRI). The relationship of structural and functional measures of brain health with cognition, and the potential modulatory role of lifestyle activities on the brain-cognition relationship were investigated.

To relate our findings to previous studies (Chan et al., 2018; Gow et al., 2017), we used the same instrument (i.e., the Lifetime of Experiences Questionnaire; LEQ [Valenzuela & Sachdev, 2007] to evaluate stimulating lifestyle activities specific to midlife, yielding two composite factors, (a) occupation and managerial responsibility, and (b) physical, social and intellectual activities. We hypothesized that midlife lifestyle factors, other than education and occupational attainment, would significantly moderate the relationship between cognition and brain health, in such a way that cognition would be de-coupled from the measures of brain health for individuals with high engagement in stimulating lifestyle activities, in line with previous research in older adults. Furthermore, we expected that such a moderation effect would be present in individuals with high CAIDE scores.

## 2 Methods

### 2.1 Participants

PREVENT-Dementia is an ongoing longitudinal multi-site research programme across the UK and Ireland, seeking to identify early biomarkers of AD and elaborate on risk-mechanism interactions for neurodegenerative diseases decades before the cardinal symptoms of dementia emerge. Its protocol has been described in detail elsewhere (Ritchie & Ritchie, 2012). In the first PREVENT programme phase, participants (40–59 years) were recruited at a single site, via the dementia register database held at the West London National Health Service (NHS) Trust, of the UK National Health Service, the Join Dementia Research website (https://www.joindementiaresearch.nihr.ac.uk/), through public presentations, social media and word of mouth. Procedures involving experiments on human participants were carried out in accord with the ethical standards of the Institutional Review Board of Imperial College London and in accord with the Helsinki Declaration of 1975. Approval for the study was granted by the NHS Research Ethics Committee London Camberwell St Giles.

Consented participants were seen at the West London NHS Trust, where they underwent a range of clinical and cognitive assessments (Ritchie & Ritchie, 2012). This subset of the ongoing PREVENT-Dementia ongoing cohort testing was particularly suitable, because cross-sectional and longitudinal data were available: 210 individuals (62 male; 148 female) were tested at baseline, and 188 (89.5%) (55 male; 133 female) tested at two-years follow-up with the same protocols (Table 1). Mild cognitive impairment and dementia were ruled out based on detailed clinical assessment on each visit.

**Table 1.**
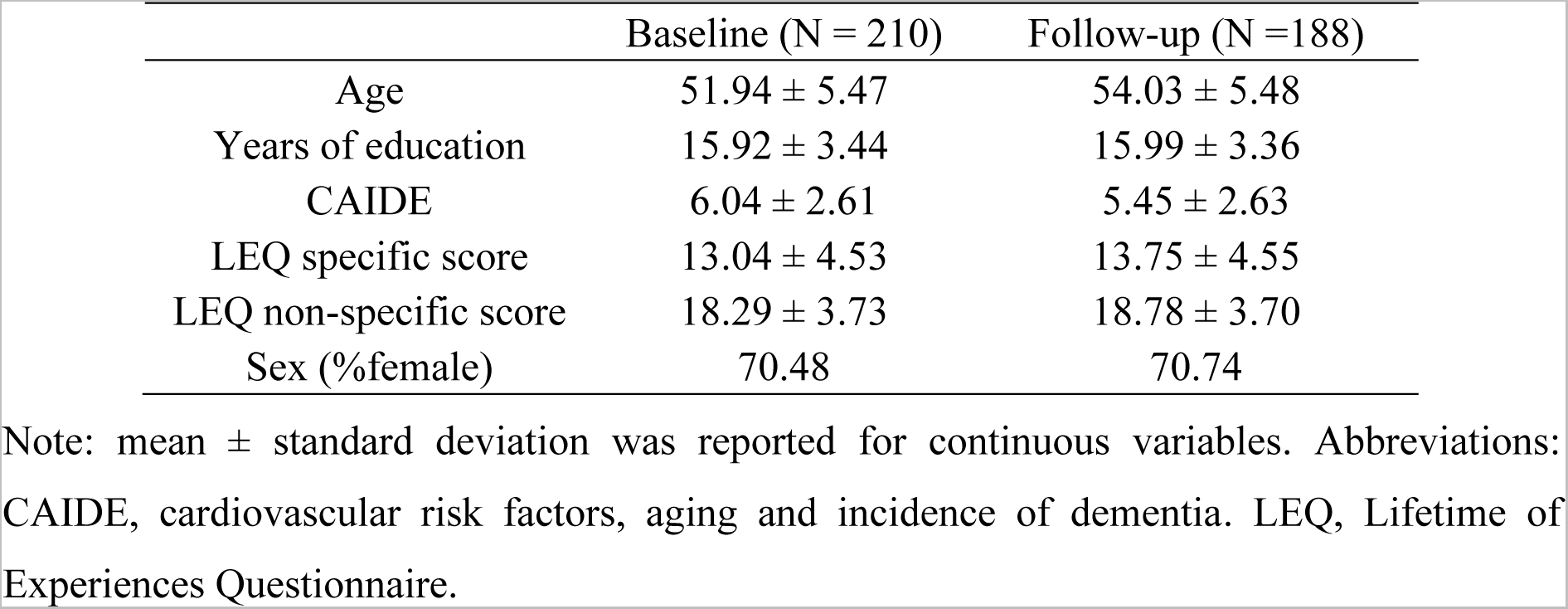
Demographic characteristics of the full cohort at baseline and follow-up

Two main analyses using different subsets of participants were conducted, as follows:

i. Subset 1: Lifestyle × brain structural health interaction on cognition. At baseline, 17 participants were excluded due to no participation or contraindications to MRI, 6 due to incidental findings on MRI scans, and 1 due to missing cognition data. At follow-up, 18 participants were excluded due to no participation or contraindications to MRI, 3 due to incidental findings on MRI scans, and 11 due to missing cognition data, resulting in N=186 at baseline and N = 156 at follow-up, for the analysis of lifestyle moderation of the association between total grey matter volume and cognition (SFigure 1).
ii. Subset 2: Lifestyle × brain functional health interaction on cognition. At baseline, 17 participants were excluded due to no participation or contraindications to MRI, 6 due to incidental findings, 20 due to incomplete brain coverage for functional brain network analysis (The subset selection based on inclusion criteria has been described in detail elsewhere [Deng et al. (2023]), and 1 due to missing cognition data. At follow-up, 19 participants were excluded due to decline or contraindications to MRI, 3 due to incidental findings, and 1 due to inadequate brain coverage, and 11 due to missing cognition data, resulting in N = 166 at baseline and N = 154 at follow-up (SFigure 1).

### 2.2 Assessments

#### 2.2.1 Dementia risk score

The CAIDE is a composite score of estimated future dementia risk that captures midlife cardiovascular risk burden (Fayosse et al., 2020; Sindi et al., 2015). It takes into account an individual’s age, sex, educational attainment, Apolipoprotein E (APOE) ε4 genotype, activity level, body mass index, cholesterol and systolic blood pressure (Kivipelto et al., 2006) and is scored on a range of 0–18. A higher score indicates a higher risk. The CAIDE dementia risk score was calculated at baseline and follow-up.

#### 2.2.2 Cognitive assessments

Cognitive function was assessed at baseline and follow-up using the COGNITO neuropsychological battery (Ritchie, 2014), which is designed to examine information processing across a wide range of cognitive functions in adults of all ages, and is not restricted to those functions usually implicated in dementia detection in older adults. Additionally, we used the Visual Short-Term Memory Binding Task (VSTMBT), which is sensitive to detecting changes in pre-symptomatic stages of AD (Parra et al., 2010), and therefore may be sensitive to subtle information processing changes due to AD risk, in asymptomatic individuals. In total, 13 measures were derived to capture multiple cognitive functions (for details see STable 1). In an independent study of this dataset (Deng et al., 2022), a dimensionality reduction method, i.e., rotated principal component analysis (rPCA) (Jolliffe and Cadima, 2016), was adopted to cluster these measures into three cognitive components (SFigure 2) in order to reduce the number of multiple comparisons, and increase the power to detect subtle effects in cognitive measures that may not exhibit behavioural variability in midlife. The rPCA was conducted using the psych package (version 2.0.12) in the R software (https://www.r-project.org/) and included the following steps: (a) component estimation by using scree plots and parallel analysis, (b) component extraction by using principal component analysis, (c) Varimax rotation to constrain the components to be uncorrelated, and (d) calculation of component scores by a regression method. In subsequent analyses, we used the three cognitive components derived from this previous work (Deng et al. 2022; SFigure 2): (i) episodic and relational memory, (ii) working and short-term (single-feature) memory, and (iii) verbal, visuospatial functions, and short-term (conjunctive) memory.

### 2.3 Measurement of lifestyle activities

The LEQ, designed to take a lifespan approach to the measurement of cognitive reserve (Stern, 2009, 2012; Stern et al., 2019) and mental activity, measures engagement in a broad range of lifestyle activities across three stages of life: young adulthood (13-29 years), midlife (30–64 years) and late life (65 years onwards). Therefore, the LEQ is preferable for looking at a midlife cohort compared to other scales that capture dementia-specific risk related to modifiable lifestyle factors (e.g., LIBRA, Schiepers et al. (2018)). The LEQ comprises sub-scores capturing “specific” activities, reflecting the primary activity undertaken in each life stage and “non-specific” activities, reflecting engagement in physical, social and intellectual activities in any stage. For the purpose of this paper, we define midlife ‘lifestyle’ as all the activities captured by the LEQ (below).

#### Midlife specific score

The midlife specific component score centres on occupation and comprises two sub-scores that measure (a) the occupational history and (b) the managerial responsibility. For the first occupational sub-score, participants were asked to record their primary occupation in each 5-year interval from age 30 to age at assessment. Each reported occupation was scored on a scale of 0-9, according to the International Standard Classification of Occupations (ISCO 08) guidelines (https://www.ilo.org/public/english/bureau/stat/isco/isco08/). This scale relates to the skill level associated with occupations, where managers score 1, professionals 2, technicians and associate professionals score 3, and so on. Participant scores were inverted and summed. The second sub-score is a measure of the managerial responsibility associated with reported occupations. If participants indicated that they were employed in a managerial capacity, the number of people that they oversaw in four of their reported occupations was documented. Managerial responsibility was scored as follows; 0 people = 8, 1-5 people = 16, 5-10 people = 24 and 10+ people = 32. The highest score is recorded as the managerial responsibility sub-score. The occupational history and managerial sub-scores were summed and multiplied by a normalization factor of 0.25. Normalization ensures that the midlife specific and non-specific scores have comparable mean values (Valenzuela & Sachdev, 2007).

#### Midlife non-specific score

The non-specific score assesses frequency of engagement in 7 activities, capturing those of a physically, socially and intellectually stimulating nature, scored on a 6-point Likert scale of frequency (never, less than monthly, monthly, fortnightly, weekly, daily). Scores range from 0 – 35, with higher scores reflecting more frequent engagement in such activities. The items included in the scale are socializing with family or friends, practicing a musical instrument, practicing an artistic pastime, engagement in physical activity that is mildly, moderately, or vigorously energetic, reading, practicing a second language and travel. The travel item asks participants if they have visited any of a list of continents between the ages of 30-54. Responses were scored on a 6-point scale as follows: none, 1-2 regions, 3-4 regions, 5 regions, 6 regions, 7 regions.

### 2.4 MRI acquisition and processing

#### Functional data acquisition and processing

MRI data were acquired using a 3T Siemens Verio scanner with a 32-channel head coil. Resting state fMRI data were acquired using a T2*-weighted EPI sequence with participants resting in the scanner with their eyes closed. A detailed description of the acquisition parameters, pre-processing of resting-state MRI data from the Imperial College London site of the PREVENT-Dementia research programme and the construction of the functional brain network, as well as the calculation of the measure of functional network segregation, i.e. the participation coefficient (Pc), can be found in Deng et al. (2023).

Briefly, resting-state fMRI data were first pre-processed using SPM12 and the AA pipeline software (Cusack et al., 2015), including motion correction, slice timing correction, co-registration of functional and structural images, normalisation to standard MNI space, spatial smoothing (with a Gaussian kernel of 6 mm full width at half maximum), band-pass filtering (0.01-0.08 Hz) and nuisance regression (24 head motion parameters) with a general linear model. We then adopted a graph-theoretic framework to guide the analysis of the functional brain network organisation. The functional brain network was constructed by defining a collection of brain nodes based on a commonly used brain parcellation scheme (Power et al., 2011) and calculating edges by Pearson correlation of pre-processed fMRI time series derived from these nodes (van den Heuvel & Hulshoff Pol, 2010). Ten subnetworks were identified in this predefined brain atlas (Power et al., 2011). We calculated the Pc of each brain node to measure the distribution of its connections among the ten subnetworks (Guimerà & Nunes Amaral, 2005; Power et al., 2013). The averaged Pc across the global brain was used to represent the segregation property of the functional brain network. Higher Pc indicates lower functional network segregation (Rubinov & Sporns, 2010).

#### Structural data acquisition and processing

T1-weighted magnetization prepared rapid gradient echo (MPRAGE) (repetition time = 2.3 s, echo time = 2.98 ms, 160 slices, flip angle = 9°, voxel size = 1 mm^3^ isotropic) scans were acquired. In an independent study by the PREVENT research team (Dounavi et al., 2022), all scans were corrected for field inhomogeneities using the Advanced Normalisation Toolbox (ANTs) N4 algorithm (Tustison et al., 2010). Freesurfer version 7.1.0 was used for data processing (Desikan et al., 2006). The recon-all pipeline was run with default settings for each participant. After recon-all, the brain masks and surfaces were inspected, and manual corrections were applied (a) in the form of erosion of non-brain voxels from the brain mask or non-WM voxels from the WM mask, (b) in the form of filling of areas where the brain was not correctly identified, or (c) with the addition of control points in cases where white matter was not successfully identified. Finally, the total grey matter volume was derived for each participant at baseline and follow-up.

### 2.5 Statistical analyses

Due to the lack of longitudinal changes over 2 years in the three investigated cognitive domains (Deng et al., 2022), the baseline and follow-up data were considered separately. Over the 2-year follow-up window, a proportion of our participants may have substantial brain health changes that are yet subthreshold to clinical manifestations. Therefore, the follow-up dataset has the potential to reveal the impact of age on the variables of interest.

The R software was used for all statistical analyses. We assessed the moderation effects of midlife lifestyle factors on the relationship between each measure of brain health and cognition using multiple linear regression, at baseline and follow-up. Each of the three cognitive scores was treated as a dependent variable, and midlife lifestyle factors (LEQ specific and non-specific scores), measures of brain health (total grey matter volume or global functional network segregation, each assessed in separate models), and their interactions were treated as independent variables. Age, sex and years of education were included as covariates in all models. For any observed moderation effect on cognition, we also tested these moderation effects for different risk groups stratified by the median of the CAIDE score. Total intracranial volume was included as a covariate for models assessing the relationship between total grey matter volume and cognition to account for individual differences in head size. To avoid multicollinearity, we mean centred continuous variables (LEQ specific and non-specific scores as well as brain health measures). A *p*-value < (0.05 / number of comparisons) was considered statistically significant to correct for multiple comparisons.

A significant interaction between a lifestyle factor and a measure of brain health on cognition would indicate that the brain-cognition coupling differs according to the levels of engagement in lifestyle activities in midlife. For any observed interactions, we plotted the regression of the brain measure on cognition for each level of lifestyle factors divided by the median value (Aiken & West, 1991), to interpret the effect. We also conducted simple slope analyses to test at which levels of engagement in lifestyle activities, a significant brain-cognition relationship was observed. Scatter plots showing these relationships were generated using unadjusted values, and full statistical details are provided in each legend for reference.

## 3 Results

### 3.1 Demographic characteristics

Demographic characteristics of the full cohort, including lifestyle activities are shown in Table 1.

### 3.2 Lifestyle factors, structural brain health and cognition

There were no significant associations between GMV × lifestyle interaction terms and cognition [episodic and relational memory (STable 2); working and short-term (single-feature) memory (STable 3); verbal, visuospatial functions, and short-term (conjunctive) memory (STable 4)], either at baseline or follow-up, after controlling for age, sex, years of education, and total intracranial volume.

### 3.3 Lifestyle factors, functional brain health and cognition

#### Episodic and Relational Memory

There were no significant associations between the global Pc × lifestyle interaction terms and cognition at either time point (STable 5).

#### Working and Short-Term (Single-Feature) Memory

Significant associations between the global Pc × LEQ non-specific score interaction and working and short-term (single-feature) memory were observed at baseline [β (SE) = 3.45 (1.54), p = 0.03], and follow-up [β (SE) = 2.91 (1.41), p = 0.04] (STable 6), but these did not survive correction for multiple comparisons.

#### Verbal and Visuospatial Functions, and Short-Term (Conjunctive) Memory

At baseline, no significant associations were observed between the global Pc × lifestyle interaction terms and verbal, visuospatial functions and short-term (conjunctive) memory (Table 2). At follow-up, there was a significant association between global Pc × non-specific LEQ and cognitive performance [β (SE) = 3.47 (1.40), p = 0.01] (Table 2). To interpret this interaction, participants were divided into low and high engagement groups based on the median of the LEQ non-specific scores, and the brain-cognition coupling for each group was examined (Figure 1). There was a significant negative relationship between global Pc and cognition for the low engagement group [β (SE) = −25.89 (8.68), p = 0.003], but no relationship for the high engagement group [β (SE) = 5.87 (8.18), p = 0.47]. For those with lower engagement in physically, socially and intellectually stimulating activities in midlife, better cognitive performance was significantly associated with lower global Pc (higher functional network segregation), as expected. By contrast, as hypothesized, cognition was independent of global Pc for those with greater engagement in lifestyle activities. In addition, we found a trend association of cognition with the global Pc × specific LEQ interaction [β (SE) = −2.92 (1.50), p = 0.05]. While this association suggests a role for occupational complexity in cognitive reserve, it is weak and needs to be further explored in future studies.

**Figure 1.**
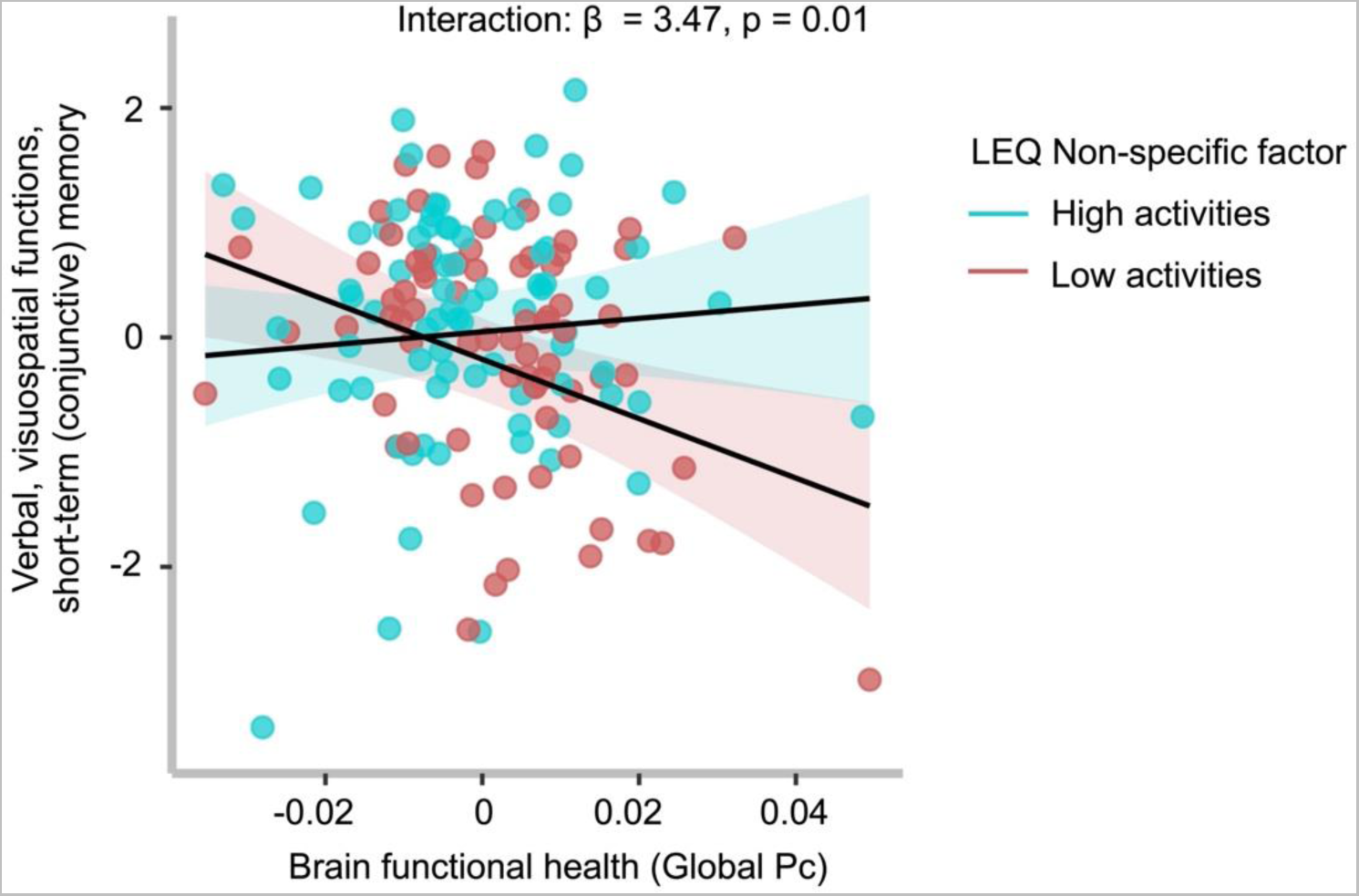
Modulation of the expected functional brain health - cognition relationship by physical, social and intellectual activities in cognitively healthy middle-aged adults. There was a significant interaction between the LEQ non-specific factor and a measure of brain functional health, global participation coefficient (Pc), on verbal, visuospatial functions, and short-term (conjunctive) memory at follow-up. On the x axis, higher scores represent lower network segregation, and on the y axis, higher scores represent better cognition. Global Pc is mean-centred. Individuals with low engagement (in salmon) in physical, social and intellectually engaging abilities showed a significant association between better cognition and greater segregation. No significant association was seen for individuals with high engagement in these activities (in cyan). The scatter plot shows unadjusted values, but the statistical significance was based on the regression analyses where we controlled for covariates.

**Table 2.** Functional brain health – Regression coefficients for Verbal and Visuospatial Functions, and Short-Term (conjunctive) Memory at Baseline & Follow-up

|  |  | Baseline |  |  | Follow-up |  |  |
| --- | --- | --- | --- | --- | --- | --- | --- |
| | | $R^2$ | F (8, 157) | $p$ | $R^2$ | F (8, 145) | $p$ |
| Model summary |  | 0.07 | 1.40 | 0.20 | 0.09 | 1.73 | 0.10 |
| DV | IV | $\beta$ | SE | $p$ | $\beta$ | SE | $p$ |
| Verbal and Visuospatial Functions, and Short-Term (conjunctive) Memory | Specific | 0.02 | 0.02 | 0.36 | 0.006 | 0.02 | 0.76 |
|  | Non-specific | 0.02 | 0.02 | 0.31 | 0.007 | 0.02 | 0.77 |
|  | Global Pc | -3.64 | 5.68 | 0.52 | -14.00 | 6.27 | 0.03 |
|  | Global Pc * Specific | 0.48 | 1.13 | 0.67 | -2.92 | 1.50 | 0.05 |
|  | Global Pc * Non-specific | -0.40 | 1.67 | 0.81 | 3.47 | 1.40 | 0.01 |
|  | Age | -0.04 | 0.02 | 0.005 | -0.02 | 0.02 | 0.30 |
|  | Sex | 0.22 | 0.17 | 0.20 | 0.24 | 0.18 | 0.19 |
|  | Years of education | -0.02 | 0.02 | 0.54 | -0.002 | 0.03 | 0.94 |

Note: unstandardized coefficients $\beta$ and standard error (SE) were reported. DV, dependent variable; IV, independent variable; Pc, participation coefficient.

We have previously shown that the above cognitive domain of “verbal, visuospatial functions, and short-term (conjunctive) memory” is negatively associated with the CAIDE score, which also captures modifiable lifestyle activities (Deng et al., 2022). Therefore, in the next analyses, we investigated whether the moderation effect of lifestyle on the relationship between this cognitive domain and brain health, is affected by the CAIDE dementia score.

For the high CAIDE group (CAIDE > 6), there was a significant relationship between global Pc × non-specific LEQ interaction and cognition [β (SE) = 4.28 (1.83), p = 0.02] (Table 3). Simple slope analyses showed that higher global Pc (lower network segregation) was significantly associated with worse cognition for individuals with low engagement [β (SE) = −33.49 (11.61), p = 0.005], but not for those with high engagement in stimulating lifestyle activities [β (SE) = −3.16 (13.22), p = 0.81] (Figure 2). This pattern of association was consistent with our hypothesis and suggests that the contribution of stimulating lifestyle activities to cognitive reserve was present only in individuals at high risk of late-life dementia as identified by the CAIDE score (CAIDE > 6). We observed a significant interaction between global Pc and the non-specific LEQ factor on cognition for the low CAIDE group [β (SE) = 5.52 (2.65), p = 0.04], but simple slope analyses showed no significant associations for each engagement group.

**Figure 2.**
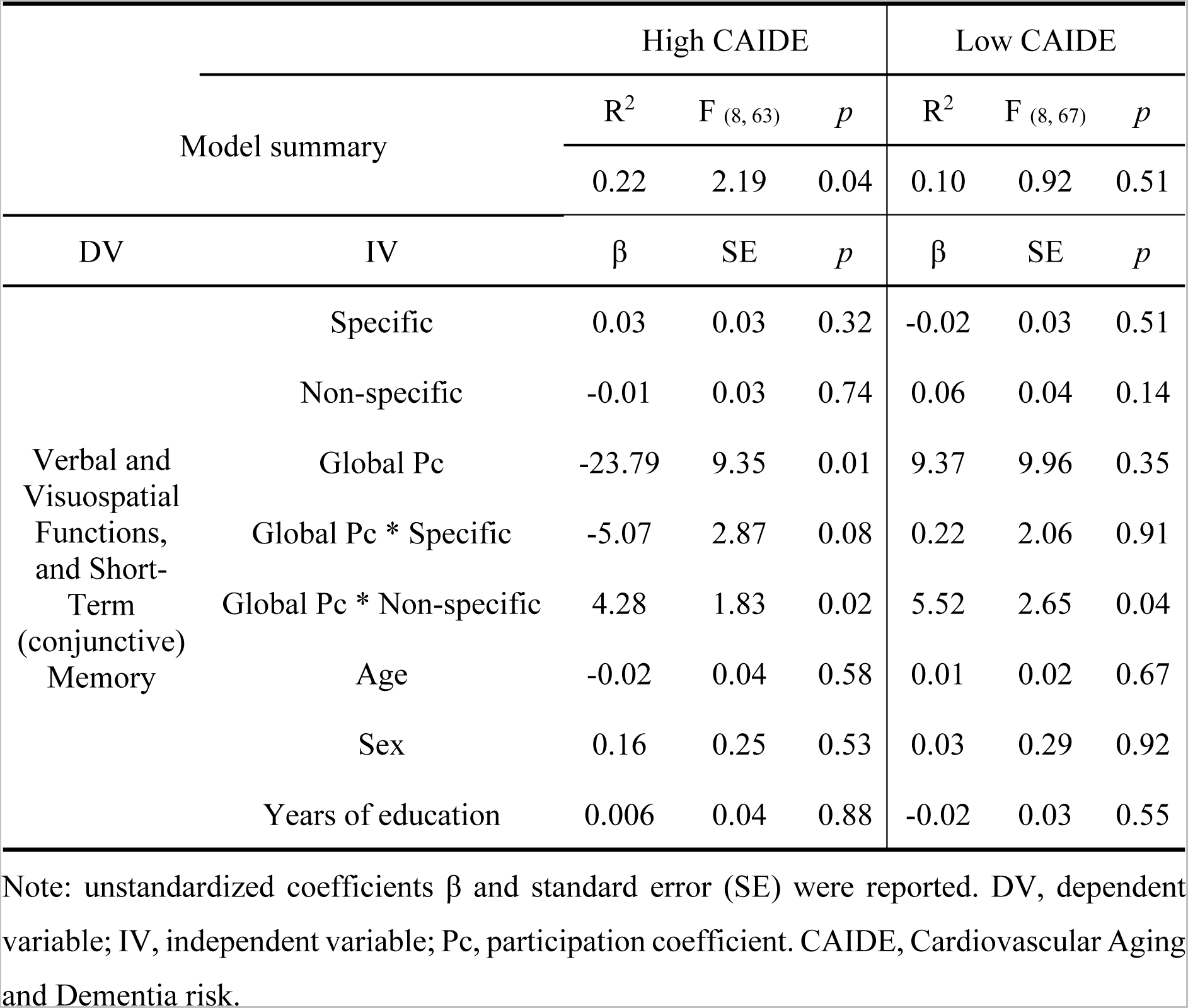
Modulation of the relationship between functional brain health and cognition relationship by physical, social and intellectual activities. On the x axis, higher scores represent lower network segregation, and on the y axis, higher scores represent better verbal, visuospatial functions, and short-term (conjunctive) memory. Global Pc is mean-centred. Expected moderation effect was present for individuals with high Cardiovascular Risk Factors, Aging, and Incidence of Dementia (CAIDE) risk score (> median = 6), but not for those with low CAIDE (≤ 6). Specifically, simple slope analyses showed a significant association between better cognition and greater functional brain network segregation only for individuals with low engagement (in red), but not for those with high engagement in these lifestyle activities (in blue). The scatter plot shows unadjusted values, but the statistical significance was based on the regression analyses where we controlled for covariates.

**Table 3.**
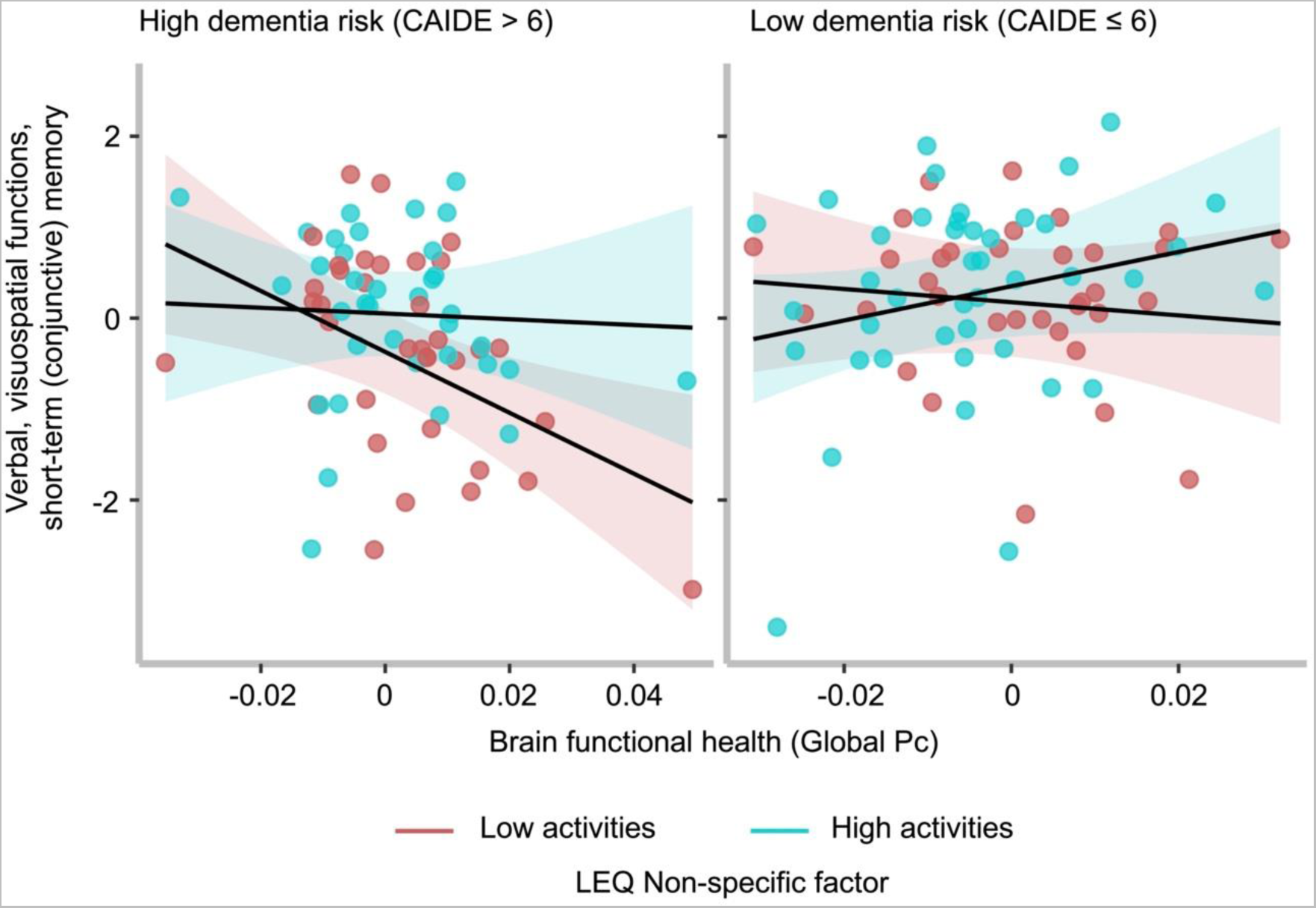
Functional brain health – Regression coefficients for Verbal and Visuospatial Functions, and Short-Term (conjunctive) Memory for different at-risk groups at follow-up.

## 4 Discussion

It is now acknowledged that Alzheimer’s disease (AD) neuropathology starts decades before the onset of clinical symptoms, but it remains unknown whether modifiable lifestyle factors can protect against these incipient AD processes, early, in mid-life. This study tested the hypothesis that lifestyle activities contribute to cognitive reserve in midlife, especially in individuals who are presently cognitively healthy, but at high risk for late-life AD (Stern et al., 2020). Consistent with the concept of cognitive reserve, we have previously shown that mid-life stimulating lifestyle activities make an independent contribution to mid-life cognition, over and above age, sex, education, and occupational attainment (Heneghan et al., 2022). A rigorous determination of cognitive reserve, however, requires further evidence that it moderates the association between a brain state and cognitive outcome (e.g., EClipSE Collaborative Members, 2010). This effect has previously been reported in older adults (Chan et al., 2018). The first novel finding of the present study was that physically, socially, and intellectually stimulating activities undertaken in mid-life significantly moderated the relationship between mid-life cognitive ability and functional brain network segregation, independent of education and occupation. The second novel finding was that this manifestation of cognitive reserve was particularly observed in middle-aged individuals with high dementia risk scores (CAIDE > 6).

We found that in individuals who had higher involvement in diverse enriching activities, cognitive ability was less dependent on functional network segregation, suggesting that they realized cognition via independent compensatory brain mechanisms bolstered by cognitive reserve. The brain health measure that was affected by cognitive reserve in this age group is functional network segregation. The brain is composed of intrinsically wired functional networks (Crossley et al., 2013; Smith et al., 2009), each corresponding to a set of distinct and tightly connected regions (Cole et al., 2014; Smith et al., 2009) and showing graded functional specialization (Sporns & Betzel, 2016; Wig, 2017). Such modular functional organisation of the brain in the form of distinct but highly interconnected networks is critical for cognition (Achard et al., 2006; Bullmore & Sporns, 2012; Chan et al., 2014). Higher functional network segregation is associated with stronger cognition across the healthy adult lifespan (Chan et al., 2014; Wig, 2017; Deng et al., 2023), and lower cognitive impairment in clinical dementia (Ewers et al. 2021). We did not find lifestyle activities to moderate the relationship between the total grey matter volume and cognition in mid-life, in contrast to previous findings in older adults (Chan et al., 2018). This is likely due to the relatively young age of this cohort, on average 30 years younger than the cohort studied in Chan et al. (2018), and 23 years prior to dementia onset (Dounavi et al., 2022), and it suggests that brain structure does not exhibit a significant impact of the avocational activities investigated herein, in mid-life.

Short-term (conjunctive) memory functions have been found impaired in the pre-symptomatic stages of AD (Parra et al., 2010). Additionally, impaired visuospatial function is one of the earliest cognitive deficits observed in AD (Parra et al., 2010) and has previously been linked to increased AD risk in this cohort (Deng et al., 2022; Ritchie et al., 2017). In similarly cognitively healthy, but older (>65 years) North American cohorts (Miron et al., 2019; Noble et al., 2010) changes in cerebrospinal fluid biomarkers related to inflammation have been associated with changes in visuospatial cognitive performance, thereby suggesting a biochemically-mediated effect of early pathology in pre-symptomatic individuals. Our results suggest that stimulating lifestyle activities may boost cognitive functions that are very vulnerable to AD risk and early AD neuropathology [Williams et al., 2020; Parra et al., 2010, Laukka et al., 2012).

The CAIDE score incorporates several modifiable risk factors (i.e., blood pressure, cholesterol, physical activity, body mass index) that reflect cardiovascular dementia risk. High CAIDE scores (> 6) has been used to select at-risk participants for a large randomised controlled trial (RCT) – the Finnish Geriatric Intervention Study to Prevent Cognitive Impairment and Disability (FINGER) – in older adults (age > 60 years). Another large RCT, the French Multidomain Alzheimer Preventive Trial (MAPT), in older adults (age > 70 years), found a significant beneficial effect of multidomain lifestyle intervention in individuals with CAIDE scores > 6 (see Kivipelto et al. (2018) for a review). Furthering these previous studies, our results suggest that stimulating lifestyle activities contribute to cognitive reserve from a much earlier life stage, when individuals are relatively young and cognitively healthy, but have risks of late-life AD. Our finding that the impact of modifiable mid-life activities was independent of educational attainment and occupational status, suggests that a public health initiative aimed at boosting cognitive reserve via enhancement of mid-life is generalizable to the entire adult population in this age range.

What might be the mechanism by which stimulating activities impact cognition and brain health in midlife? Models of Alzheimer’ disease delineated by neuropathological staging (Braak & Braak, 1991) place the locus coeruleus (LC), a small nucleus in the pontine tegmentum region of the brainstem, at the pathogenesis of AD from late young adulthood and early midlife (30-40 years) (Arnsten et al., 2021; Jacobs et al., 2021; Márquez & Yassa, 2019). The LC is responsible for the production of the neurotransmitter noradrenaline (NA) (Amaral & Sinnamon, 1977), a major driver of the brain’s arousal system, which strongly modulates high-order cognition. Novelty provides a key trigger for arousal, and thus LC–NA activity occurs strongly in response to novel stimuli. Studies (Robertson, 2013, 2014) have suggested that environmental enrichment through lifestyle stimulating activities, such as those investigated in this study, upregulates the noradrenergic system, which is otherwise depleted with age (Liu et al., 2020) and AD pathology (Jacobs et al., 2021), leading to compensatory brain mechanisms for cognitive function, such as the strengthening of the fronto-parietal brain and other large-scale brain networks.

Several limitations should be considered. First, as education is strongly positively linked to IQ (Ritchie & Tucker-Drob, 2018) and our cohort was highly educated, the question of reverse causation arises (Borgeest et al., 2020; Chan et al., 2018; Gow et al., 2017). This points to the possibility that cognitive abilities may determine engagement in stimulating activities, rather than the inverse. However, the effect on midlife verbal, visuospatial functions, and short-term memory is independent of the total years of education, which shows that education does not directly drive the variability of this domain. Second, a longitudinal effect of lifestyle on cognition was not obtained in this study, due to the lack of longitudinal cognitive change, as previously reported in this cohort (Deng et al., 2022). Previous studies from this cohort have shown only subtle changes over the two-year period in brain measures but not in cognition (Dounavi et al., 2021; Low et al., 2021), possibly due to the relatively young age range and the short follow-up window (Ritchie et al., 2018). Future studies following this cohort for a longer period of time, i.e., 5 years, are underway and will be important to address the longitudinal impact of lifestyle activities in this cognitively healthy middle-aged cohort, particularly in those at risk of late-life Alzheimer’s disease. Third, we caution that the interpretability of the effect of lifestyle activities on each individual cognitive function is limited by their composite assessment in this study and requires individuation in future studies with a longer longitudinal follow-up window.

In conclusion, our findings suggest that stimulating lifestyle activities in mid-life contribute to cognitive reserve and may offset cognitive decrements due to AD risk in this young age group. The modifiability of these activities supports their targeting by public health initiatives aimed at dementia prevention.

## Data Availability

All data produced in the present study are available upon reasonable request to the authors

## Acknowledgement

F.D. was funded by the Provost PhD Award Scheme from Trinity College Dublin, to L.N. L.N. was also funded by a L’Oréal-UNESCO for Women in Science International Rising Talent Award, the Welcome Trust Institutional Strategic Support grant, and the Global Brain Health Institute Project Grant.

The PREVENT-Dementia study is supported by the UK Alzheimer’s Society (grant numbers 178 and 264), the US Alzheimer’s Association (grant number TriBEKa-17-519007) and philanthropic donations.

We thank all PREVENT-Dementia participants for their enthusiastic participation in this study. We also thank the Research Delivery service at West London NHS Trust and the Wolfson Clinical Imaging Facility at Imperial College London for their support in running the study.

