## Supplementary Information for "Lifestyle activities in mid-life contribute to cognitive reserve in middle-aged individuals at risk for late-life Alzheimer’s disease, independent of education and occupation"

**Title**

^*^ **Corresponding author:**

Lorina Naci

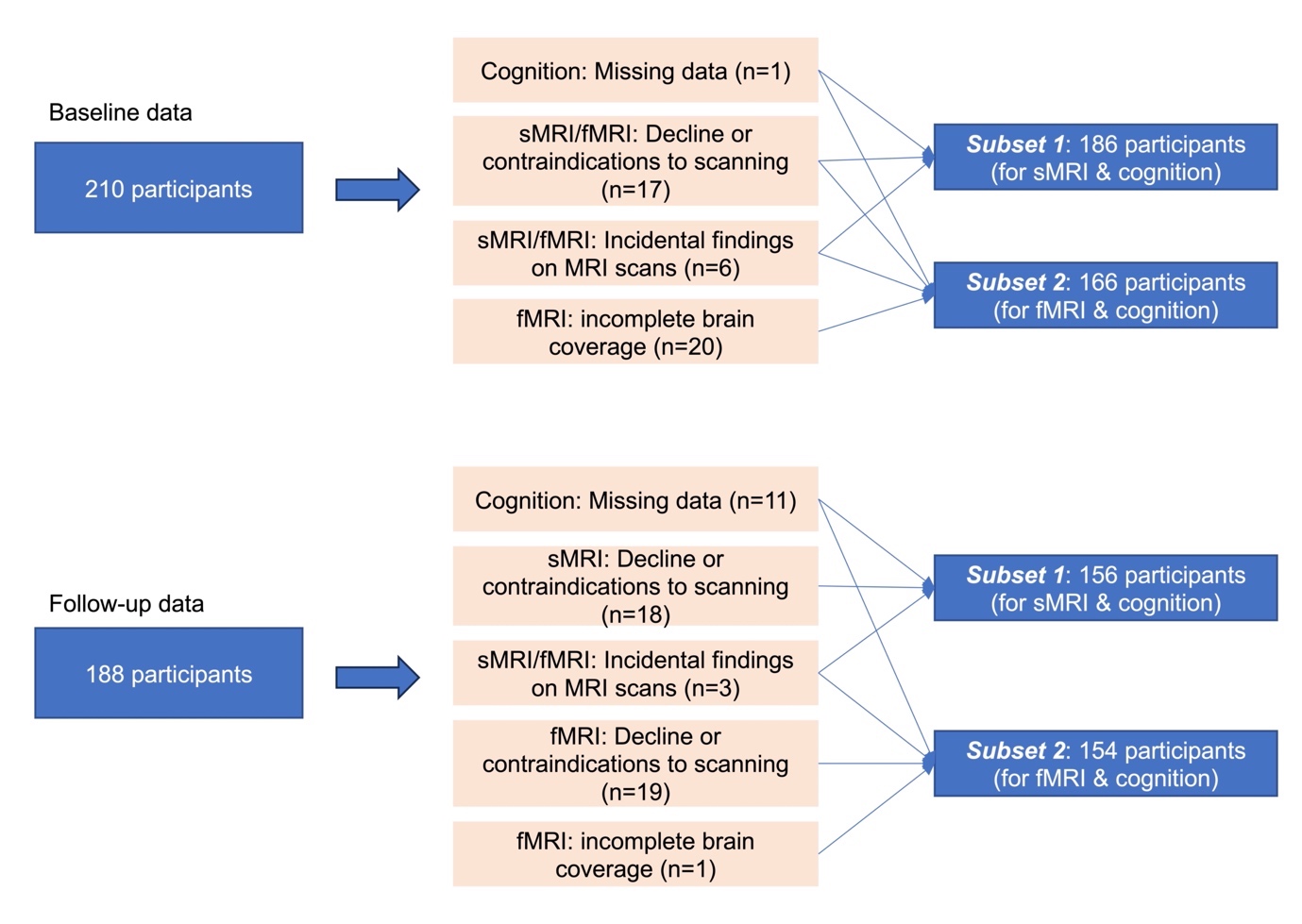

SFigure 1 Participant inclusions for different analyses. Abbreviations: sMRI, structural magnetic resonance imaging; fMRI, functional magnetic resonance imaging.

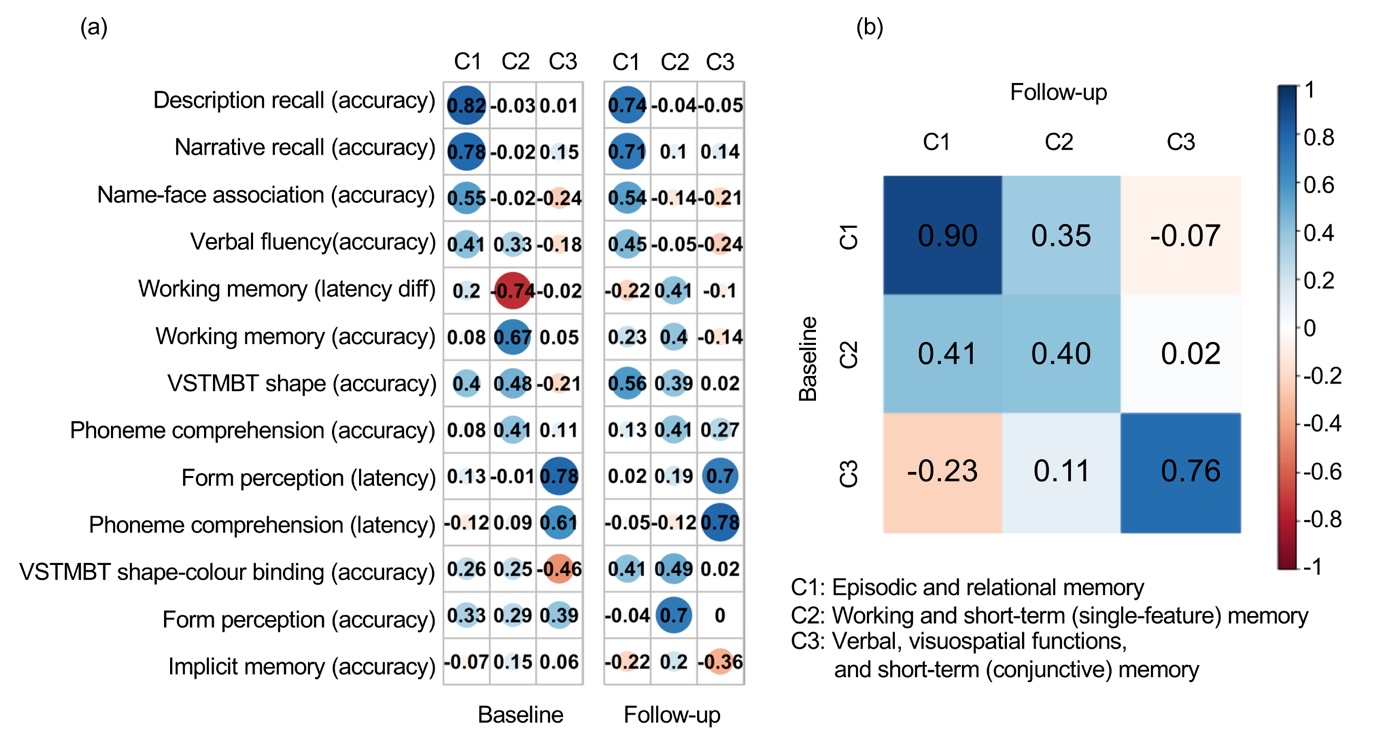

SFigure 2 Interpretation of data-derived cognitive domains. Component (C) coefficients and Tucker's congruence coefficients between (a) baseline and (b) follow-up, are shown. (a) The coefficients for 13 original cognitive measures (in rows) on the three components (in columns). A larger absolute coefficient (darker colors and larger solid circles) represents a closer relationship between the cognitive measure and corresponding component. Cool/warm colors represent the positive/negative relationships between cognitive measures and components. (b) Similarities among the components across baseline (in rows) and follow-up (in columns) were measured by Tucker's congruence coefficients. The diagonal values represented the similarities for each of the components with itself across time, and off diagonal values indicated the similarities for each of the components with the other two components across time. Cool/warm colors represent the positive/negative relationships with darker colors representing correlation magnitude, as shown in the color-bar scale. Abbreviations: VSTMBT, visual short-term memory binding test; diff, difference. This figure is adapted from Deng et al., 2023.

| STable 1. Description of cognitive tasks and measures | | | | |
| --- | --- | --- | --- | --- |
| Cognitive Tasks | Measures | Task Description | Descriptive Statistics (N=210, baseline) | Descriptive Statistics (N=188, follow-up) |
| Working memory | Total number of correct answers | Dual task: The subject must locate the targeted shapes and count the sounds. | Mean = 9.75 SD = 0.54 Range = 3.00 Missingness = 0 | Mean = 9.85 SD = 0.51 Range = 3.00 Missingness = 4 |
|  | Mean time (ms) difference in milliseconds between the dual task and a simple form recognition task | Dual task: The subject must locate the targeted shapes and count the sounds.  Simple task: Subject must locate the targeted shapes only. | Mean = 223.09 SD = 3175.07 Range = 18238.00 Missingness = 0 | Mean = -55.47 SD = 2919.28 Range = 17750.00 Missingness = 4 |
| Narrative recall | Total number of correct answers | The subject must recall a series of elements which have a logical sequence (a short story). | Mean = 13.25 SD = 4.73 Range = 23.00 Missingness = 0 | Mean = 15.03 SD = 4.30 Range = 23.00 Missingness = 4 |
| Description recall | Total number of correct answers | The subject must recall a series of elements which have a visual sequence (a short description). | Mean = 12.63 SD = 4.34 Range = 20.00 Missingness = 0 | Mean = 13.39 SD = 4.73 Range = 25.00 Missingness = 4 |
| Implicit memory | Difference the number of names never seen and the number of names already learned | The subject must recognize as soon as possible a name which is constructed progressively on the screen. | Mean = 1.05 SD = 0.83 Range = 8.20 Missingness = 0 | Mean = 1.11 SD = 0.69 Range = 5.00 Missingness = 4 |
| Name-face association | The number of correctly recognized faces and their corresponding names | The subject must decide whether a face on the screen appeared before and if yes, what the person's name is. | Mean = 5.37 SD = 2.21 Range = 9.00 Missingness = 0 | Mean = 5.86 SD = 2.02 Range = 9.00 Missingness = 4 |

| STable 1. Description of cognitive tasks and measures (continued) | | | | |
| --- | --- | --- | --- | --- |
| Form matching | Total number of correct answers | The subject must discriminate form and line orientation by matching a sample complex figure to one of six figures. Distractor figures are designed to detect visuospatial field neglect and difficulties with line orientation. | Mean = 6.45 SD = 1.12 Range = 7.00 Missingness = 0 | Mean = 6.43 SD = 1.01 Range = 4.00 Missingness = 4 |
|  | Mean time (ms) for correct answers |  | Mean = 5841.64 SD = 1425.88 Range = 7596.00 Missingness = 0 | Mean = 6071.80 SD = 1567.05 Range = 8116.00 Missingness = 4 |
| Phoneme comprehension | Total number of correct answers | The subject must choose an object illustrating a presented word among 6 objects which include shape, phonetic and semantic distractors. | Mean = 8.62 SD = 0.57 Range = 3.00 Missingness = 0 | Mean = 8.63 SD = 0.53 Range = 2.00 Missingness = 4 |
|  | Mean time (ms) for correct answers |  | Mean = 1585.71 SD = 304.24 Range = 1654.00 Missingness = 0 | Mean = 1508.96 SD = 272.53 Range = 1724.00 Missingness = 4 |
| Verbal fluency | Total number of correct answers | The subject must name all the words they can think of within one minute based on semantic and phonetic cues. | Mean = 28.39 SD = 6.58 Range = 32.00 Missingness = 0 | Mean = 29.59 SD = 7.12 Range = 43.00 Missingness = 4 |
| VSTMBT-shape only | Total number of correct answers | The subject must recall stimuli that were shapes after a short period of retention. | Mean = 0.85 SD = 0.14 Range = 0.88 Missingness = 2 | Mean = 0.86 SD = 0.17 Range = 2.31 Missingness = 10 |
| VSTMBT-shape colour binding | Total number of correct answers | The subject must recall stimuli that were combinations of shapes and colours after a short period of retention. | Mean = 0.52 SD = 0.20 Range = 1.19 Missingness = 2 | Mean = 0.54 SD = 0.24 Range = 2.19 Missingness = 11 |
| Key: VSTMBT, visual short-term memory binding test; SD, standard deviation | | | | |

STable 2 Brain structural health - Regression coefficients for Episodic and Relational Memory at Baseline & Follow-up

|  |  | Baseline | | | Follow-up | | |
| --- | --- | --- | --- | --- | --- | --- | --- |
| Model summary | | R^2^ | F _(9, 176)_ | *p* | R^2^ | F _(9, 146)_ | *p* |
|  |  | 0.19 | 4.50 | <0.0001 | 0.24 | 5.16 | <0.0001 |
| DV | IV | β | SE | *p* | β | SE | *p* |
| Episodic and Relational Memory | Specific | -0.02 | 0.02 | 0.20 | -0.03 | 0.02 | 0.16 |
|  | Non-specific | 0.04 | 0.02 | 0.06 | 0.04 | 0.02 | 0.05 |
|  | TGMV | -3.36e-06 | 3.00e-06 | 0.26 | -2.56e-06 | 3.31e-06 | 0.44 |
|  | TGMV * Specific | -1.13e-07 | 2.89e-07 | 0.70 | -1.35e-07 | 3.69e-07 | 0.72 |
|  | TGMV * Non-specific | 5.67e-08 | 4.59e-07 | 0.90 | 1.41e-08 | 4.79e-07 | 0.98 |
|  | Age | -0.01 | 0.01 | 0.33 | -0.02 | 0.02 | 0.27 |
|  | Sex | 0.21 | 0.19 | 0.27 | 0.21 | 0.21 | 0.32 |
|  | Years of education | 0.10 | 0.02 | <0.0001 | 0.12 | 0.02 | <0.0001 |
|  | TICV | 1.65e-06 | 1.00e-06 | 0.10 | 1.33e-06 | 1.13e-06 | 0.24 |

Note: unstandardized coefficients β and standard error (SE) were reported. DV, dependent variable; IV, independent variable; TGMV, total grey matter volume; TICV, total intracranial volume

STable 3 Brain structural health - Regression coefficients for Working and Short-Term (single-feature) Memory at Baseline & Follow-up

|  |  | Baseline | | | Follow-up | | |
| --- | --- | --- | --- | --- | --- | --- | --- |
| Model summary | | R^2^ | F _(9, 176)_ | *p* | R^2^ | F _(9, 146)_ | *p* |
|  |  | 0.05 | 1.04 | 0.41 | 0.07 | 1.29 | 0.25 |
| DV | IV | β | SE | *p* | β | SE | *p* |
| Working and Short-Term (single-feature) Memory | Specific | 0.02 | 0.02 | 0.43 | 0.02 | 0.02 | 0.33 |
|  | Non-specific | 0.007 | 0.02 | 0.73 | -0.04 | 0.02 | 0.13 |
|  | TGMV | 3.44e-07 | 3.28e-06 | 0.92 | -8.17e-07 | 3.61e-06 | 0.82 |
|  | TGMV * Specific | 6.06e-08 | 3.16e-07 | 0.85 | 7.33e-07 | 4.02e-07 | 0.07 |
|  | TGMV * Non-specific | 3.32e-08 | 5.01e-07 | 0.95 | -2.32e-07 | 5.22e-07 | 0.66 |
|  | Age | -0.02 | 0.02 | 0.24 | -0.02 | 0.02 | 0.21 |
|  | Sex | -0.11 | 0.20 | 0.60 | -0.34 | 0.23 | 0.13 |
|  | Years of education | 0.04 | 0.02 | 0.10 | 0.005 | 0.03 | 0.84 |
|  | TICV | 3.32e-07 | 1.10e-06 | 0.76 | 2.36e-07 | 1.23e-06 | 0.85 |

Note: unstandardized coefficients β and standard error (SE) were reported. DV, dependent variable; IV, independent variable; TGMV, total grey matter volume; TICV, total intracranial volume

STable 4 Brain structural health - Regression coefficients for Verbal, Visuospatial Functions, and Short-Term (conjunctive) Memory at Baseline & Follow-up

|  |  | Baseline | | | Follow-up | | |
| --- | --- | --- | --- | --- | --- | --- | --- |
| Model summary | | R^2^ | F _(9, 176)_ | *p* | R^2^ | F _(9, 146)_ | *p* |
|  |  | 0.08 | 1.71 | 0.09 | 0.05 | 0.86 | 0.57 |
| DV | IV | β | SE | *p* | β | SE | *p* |
| Verbal, Visuospatial Functions, and Short-Term (conjunctive) Memory | Specific | 0.01 | 0.02 | 0.55 | -0.001 | 0.02 | 0.95 |
|  | Non-specific | 0.04 | 0.02 | 0.10 | 0.02 | 0.02 | 0.49 |
|  | TGMV | -7.56e-07 | 3.32e-06 | 0.82 | 4.44e-06 | 3.61e-06 | 0.22 |
|  | TGMV * Specific | 7.78e-08 | 3.19e-07 | 0.81 | -5.38e-07 | 4.02e-07 | 0.18 |
|  | TGMV * Non-specific | -7.99e-07 | 5.06e-07 | 0.12 | 7.99e-07 | 5.22e-07 | 0.13 |
|  | Age | -0.05 | 0.02 | 0.004 | -0.02 | 0.02 | 0.26 |
|  | Sex | 0.22 | 0.21 | 0.28 | 0.27 | 0.23 | 0.23 |
|  | Years of education | -0.03 | 0.02 | 0.26 | -0.002 | 0.03 | 0.94 |
|  | TICV | 3.72e-07 | 1.11e-06 | 0.74 | -1.04e-06 | 1.23e-06 | 0.40 |

Note: unstandardized coefficients β and standard error (SE) were reported. DV, dependent variable; IV, independent variable; TGMV, total grey matter volume; TICV, total intracranial volume

STable 5 Brain functional health - Regression coefficients for Episodic and Relational Memory at Baseline & Follow-up

|  |  | Baseline | | | Follow-up | | |
| --- | --- | --- | --- | --- | --- | --- | --- |
| Model summary | | R^2^ | F _(8, 157)_ | *p* | R^2^ | F _(8, 145)_ | *p* |
|  |  | 0.21 | 5.08 | <0.0001 | 0.25 | 5.94 | <0.0001 |
| DV | IV | β | SE | *p* | β | SE | *p* |
| Episodic and Relational Memory | Specific | -0.02 | 0.02 | 0.33 | -0.02 | 0.02 | 0.19 |
|  | Non-specific | 0.03 | 0.02 | 0.20 | 0.04 | 0.02 | 0.04 |
|  | Global Pc | -13.10 | 5.04 | 0.01 | 0.51 | 5.80 | 0.93 |
|  | Global Pc * Specific | 0.75 | 1.00 | 0.45 | 0.29 | 1.39 | 0.84 |
|  | Global Pc * Non-specific | -1.93 | 1.48 | 0.19 | 1.84 | 1.30 | 0.16 |
|  | Age | -0.01 | 0.01 | 0.39 | -0.02 | 0.02 | 0.29 |
|  | Sex | 0.20 | 0.15 | 0.20 | 0.19 | 0.17 | 0.27 |
|  | Years of education | 0.10 | 0.02 | <0.0001 | 0.12 | 0.02 | <0.0001 |

Note: unstandardized coefficients β and standard error (SE) were reported. DV, dependent variable; IV, independent variable; Pc, participation coefficient.

STable 6 Brain functional health - Regression coefficients for Working and Short-Term (single-feature) Memory at Baseline & Follow-up

|  |  | Baseline | | | Follow-up | | |
| --- | --- | --- | --- | --- | --- | --- | --- |
| Model summary | | R^2^ | F _(8, 157)_ | *p* | R^2^ | F _(8, 145)_ | *p* |
|  |  | 0.08 | 1.74 | 0.09 | 0.08 | 1.59 | 0.13 |
| DV | IV | β | SE | *p* | β | SE | *p* |
| Working and Short-Term (single-feature) Memory | Specific | 0.01 | 0.02 | 0.54 | 0.01 | 0.02 | 0.56 |
|  | Non-specific | 0.02 | 0.02 | 0.32 | -0.04 | 0.02 | 0.11 |
|  | Global Pc | 6.41 | 5.21 | 0.22 | -7.60 | 6.31 | 0.23 |
|  | Global Pc * Specific | -0.14 | 1.04 | 0.89 | -1.52 | 1.51 | 0.32 |
|  | Global Pc * Non-specific | 3.45 | 1.54 | 0.03 | 2.91 | 1.41 | 0.04 |
|  | Age | -0.02 | 0.01 | 0.30 | -0.01 | 0.02 | 0.45 |
|  | Sex | -0.20 | 0.16 | 0.21 | -0.32 | 0.18 | 0.08 |
|  | Years of education | 0.03 | 0.02 | 0.27 | 0.003 | 0.03 | 0.91 |

Note: unstandardized coefficients β and standard error (SE) were reported. DV, dependent variable; IV, independent variable; Pc, participation coefficient.
